# Incidence of Creutzfeldt-Jakob Disease in the United States from 2000 to 2019

**DOI:** 10.1101/2024.05.14.24307294

**Authors:** Alison Seitz, Cenai Zhang, Babak B. Navi, Hooman Kamel, Alexander E. Merkler

**Affiliations:** Clinical and Translational Neuroscience Unit, Feil Family Brain and Mind Research Institute and Department of Neurology, Weill Cornell Medical College, New York, NY

**Keywords:** Creutzfeldt-Jakob Disease, prion disease, rapidly progressive dementia, epidemiological study

## Abstract

**Background and purpose:** To test the hypothesis that the incidence of Creutzfeldt-Jakob disease (CJD) has remained constant, we calculated the rate of hospitalizations for CJD in the United States using the National Inpatient Sample (NIS) from 2000 to 2019.

**Methods:** We used ICD-9 and ICD-10 codes to identify people hospitalized with presumed CJD in the National Inpatient Sample (NIS) from 2000 to 2019. Survey weights were used to calculate nationally representative estimates. We used 2000 census data to calculate age-adjusted standardized rates of CJD hospitalizations by sex and race-ethnicity and then used Joinpoint regression to evaluate changes in those rates.

**Results:** From 2000 to 2019, there were 11,064 admissions for CJD across the U.S. Across this period, the age-adjusted rate of CJD-related hospitalizations increased significantly from 1.25 (95% CI, 1.25-1.26) to 1.98 (95% CI, 1.98-1.99) per million U.S. adults per year, with a significant annual percentage change between 2004 and 2013 of 7.6% (95% CI, 4.4%-10.9%).

**Conclusions:** The incidence of CJD increased in the United States from 2000 to 2019, with a significant increase specifically between 2004 and 2013, though the overall case rate remains low.

---

Creutzfeldt-Jakob disease (CJD) is a rapidly progressive dementia caused by transmissible misfolded proteinaceous infectious particles (prions). Patients with CJD typically present with behavioral abnormalities, ataxia, extrapyramidal features, and myoclonus. CJD is always fatal, usually within a year of symptom onset.^1–3^ Sporadic CJD accounts for more than 85% of cases, and genetic forms account for about 10%.^1,4–6^ Growth hormone treatment or dura mater grafts resulted in iatrogenic CJD cases mostly in the 1990s, and the consumption of cattle infected with bovine spongiform encephalopathy led to variant CJD in the 1990s and early 2000s.^1,5,6^ Many countries have CJD surveillance systems, but these may fail to capture all suspected cases of CJD. For example, compared with hospital discharge records, the Belgian surveillance system captured only about 60% of fatalities from suspected CJD.^7^ While some studies indicate that the overall number of cases of CJD is increasing, it is unclear if that increase is due to population growth, the aging of the population, or specific increases in sex or race-ethnicity subgroups.^3,6,8–11^ Given these uncertainties, we sought to determine whether the incidence of CJD had changed in the United States from 2000 to 2019.

## Methods

### Study Design

This is a cross-sectional study using the National Inpatient Sample (NIS), a nationally representative, deidentified survey performed by the Agency for Healthcare Research and Quality. The largest all-payer medical sample in the USA, the NIS includes data on approximately 8 million inpatient hospitalizations each year, representing a 20% stratified sample of all nonfederal US hospitals. Each de-identified discharge record includes one primary diagnosis and up to 14 secondary diagnoses, using the International Classification of Diseases, Ninth and Tenth Editions (ICD-9 and ICD-10). The NIS also provides information regarding patient demographics, comorbidities, procedures, case-severity measures, and hospital characteristics.^12,13^ This study was approved by the institutional review board at Weill Cornell Medical College and the need for obtaining informed consent was waived.

### Population and Measurements

We included all adult hospitalizations from 2000 to 2019 with a diagnosis of CJD, defined by the ICD-9 discharge diagnosis codes 046.1 (which includes 046.11 and 046.19) or the ICD-10 code A81.0 (which includes A81.00, A81.01, and A81.09) in any diagnosis code position. Prior studies have used the ICD-9 code 046.1^2,8,14–16^ and the ICD-10 code A81.0^8,11,14,16,17^ from cause of death or hospital discharge datasets to track CJD diagnosis in several countries.

### Statistical Analysis

We used sampling weights provided by the NIS to calculate nationally representative estimates, and population estimates from the US Census to calculate hospitalization rates per 1 million U.S. adults per year.^12,18,19^ Because of the NIS sample redesign, we used the updated trend weights for 2000 to 2011 and the original discharge weights for 2012 to 2019. Age-adjusted rates were calculated using 2000 U.S. census as the standard population. We conducted statistical analysis using STATA (version 15.1, College Station, Texas) and Joinpoint Regression Software (version 4.9.1, National Cancer Institute).^18,20^ The joinpoint regression model assumes that data may be divided into segments and each segment may have its own trend. Segments are connected at joinpoints. The Joinpoint Regression Software determined the number of significant joinpoints by performing a series of permutation tests and used Monte Carlo methods to calculate a p-value for each test. After the number of joinpoints was determined, the annual percentage change was calculated for each of the detected time segments. The same procedure was used in subgroup analyses to detect sex and race differences in the trend. All trend analyses were 2-sided tests, and statistical significance was defined as *p*<0.05.

## Results

From 2000 to 2019, there were 11,064 admissions for CJD across the U.S. Across this period, age-adjusted rate of CJD-related hospitalizations increased significantly from 1.25 (95% CI, 1.25-1.26) to 1.98 (95% CI, 1.98-1.99) per million adults per year, with a significant annual percentage change between 2004 and 2013 of 7.6% (95% CI, 4.4%-10.9%; p<.001; Figure 1). The annual percentage change was -3.3% (CI -13.9%-8.7%) from 2000 to 2004 and -1.0 (CI -4.7%-2.8%) from 2013 to 2019. Subgroup analyses by sex and race-ethnicity showed similar results (Supplemental Materials).

**Figure 1:**
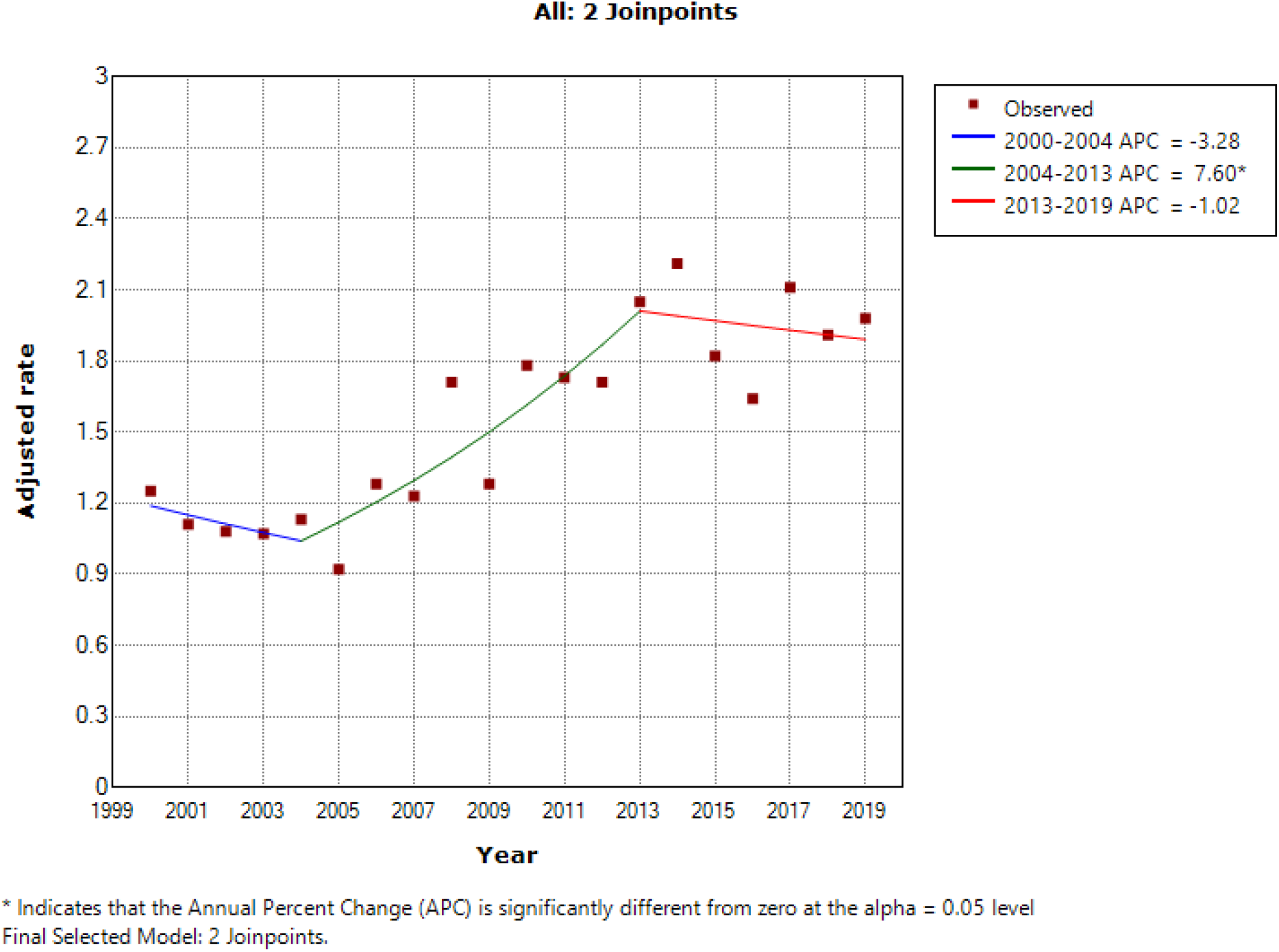
Trend in the rate of age-adjusted hospitalizations for Creutzfeldt-Jakob Disease per million persons by year

## Discussion

We found that total age-adjusted CJD-related hospitalizations per year in the United States increased between 2000 and 2019.

### Prior studies of CJD incidence

Our results should be taken in the context of prior work evaluating the incidence of CJD. Several countries have tracked CJD incidence since 1970. A 2020 metanalysis using data predominantly from the CJD International Surveillance Network EuroCJD and the United Kingdom National CJD Research and Surveillance Unit estimated the incidence of CJD to be around 1–2 cases per million per year from 2005 to 2018. The metanalysis noted a steady increase in sporadic CJD cases between 1996 and 2018, with continually low cases of iatrogenic, genetic, and variant forms of CJD.^3^ The United States Centers for Disease Control and Prevention tracks CJD deaths using a combination of ICD codes with other surveillance mechanisms, and reports an age-adjusted rate of roughly 1 to 1.5 cases per million person-years from 1979 to 2020.^21^ Our study and another recent study, which reported an age-adjusted incidence of 1.4 per million person-years,^11^ detected an increasing age-adjusted incidence of CJD per million person-years. Other studies have found age-adjusted mortality from CJD ranging from 0.19 to 2.29 during the years 1979 to 2018 in Europe, Australia, Canada, Japan, and the United States,^2,4–9,14,22–24^ with some studies reporting an increase.^6,8,9^ In this context, our study provides the novel finding that, although the incidence of CJD did increase for approximately a decade starting in 2004, the incidence appears to have plateaued starting in 2013.

### Limitations

Our study has several limitations. First, as we used the NIS to capture cases of CJD, patients who never presented to the hospital were not included. However, as the majority of patients with CJD are hospitalized at some point in time, we think this is unlikely to substantively affect our results.^8^ Second, ICD codes are an imperfect surveillance tool. In Massachusetts from 2000 to 2008, a death certificate diagnosis of CJD, which closely matched hospital discharge dataset diagnosis code 046.1, was 71% sensitive and 75% specific for pathologically confirmed CJD.^25^ In the United States between 2003 and 2005, 90.7% of suspected prion disease cases for which physicians obtained neuropathologic results were confirmed.^11^ Similarly, in Australia between 1970 and 1999, the code 046.1 on a death certificate had a sensitivity of 83.0%, and a 11.5% false-positive rate for CJD, yielding a positive predictive value of 88.5%.^26^ This is likely because histopathological examination, usually following autopsy rather than diagnostic brain biopsy, is the only way to establish a definite diagnosis of sporadic CJD, and not all CJD cases are referred for autopsy.^27^ Instead, CJD is usually diagnosed using a combination of brain MRI with abnormal cortical or deep nuclei gray matter on the diffusion-weighted imaging (DWI) and apparent diffusion coefficient (ADC) sequences, which is more than 90% sensitive and specific for sporadic CJD, combined with markers for neuronal injury, and, in late stages of the disease, EEG.^1,28^ These diagnostic tools are relatively good but imperfect. In a study in Japan, neuropathologic examination during autopsy confirmed 53 or 60 cases of suspected CJD.^29^ Similarly, in a study of ten countries’ CJD surveillance systems from 1993 to 2006, 73% of all suspected prion disease cases that underwent neuropathological examination were classified as definite cases.^4^ The imperfection of the ICD-9 and ICD-10 codes for CJD is a reflection of the difficulty in diagnosing CJD without biopsy. Lastly, our findings may be affected by changes in hospital coding practices over time, particularly around the transition to ICD-10 in late 2015. However, while this transition may have affected the absolute rates, it seems unlikely to explain the change in trend.

In conclusion, the incidence of CJD increased in the United States from 2000 to 2019, with a significant increase specifically between 2004 and 2013, though the overall case rate remains low.

## Data Availability

The statistical analysis that supports the findings of this study are available from the corresponding author upon reasonable request. The data that support the findings of this study may be requested from the Agency for Healthcare Research and Quality.

## Acknowledgements

None.

### Appendix 1: Authors

| **Name** | **Location** | **Contribution** |
| --- | --- | --- |
| Alison Seitz, MD | Weill Cornell Medical College, New York | Drafted and revised the manuscript for content, interpreted data for the work, and prepared the final format for submission |
| Cenai Zhang, MS | Weill Cornell Medical College, New York | Performed statistical analysis and performed critical revision of the manuscript |
| Babak B. Navi, MD, MS | Weill Cornell Medical College, New York | Performed critical revision of the manuscript |
| Hooman Kamel, MD, MS | Weill Cornell Medical College, New York | Performed critical revision of the manuscript |
| Alexander E. Merkler, MD | Weill Cornell Medical College, New York | Performed statistical analysis, drafted and revised the manuscript for content, interpreted data for the work, and provided final approval of the version to be submitted |

## References

1. Geschwind MD. Prion Diseases. CONTINUUM: Lifelong Learning in Neurology. 2015;21(6):1612. doi:10.1212/CON.0000000000000251

2. Gibbons RV, Holman RC, Belay ED, Schonberger LB. Creutzfeldt-Jakob disease in the United States: 1979-1998. Jama. 2000;284(18):2322–2323.

3. Uttley L, Carroll C, Wong R, Hilton DA, Stevenson M. Creutzfeldt-Jakob disease: a systematic review of global incidence, prevalence, infectivity, and incubation. The Lancet Infectious Diseases. 2020;20(1):e2–e10. doi:10.1016/S1473-3099(19)30615-2

4. Klug GMJA, Wand H, Simpson M, et al. Intensity of human prion disease surveillance predicts observed disease incidence. J Neurol Neurosurg Psychiatry. 2013;84(12):1372–1377. doi:10.1136/jnnp-2012-304820

5. Brandel JP, Peckeu L, Haïk S. The French surveillance network of Creutzfeldt–Jakob disease. Epidemiological data in France and worldwide. Transfusion Clinique et Biologique. 2013;20(4):395–397. doi:10.1016/j.tracli.2013.02.029

6. Ladogana A, Puopolo M, Croes E, et al. Mortality from Creutzfeldt–Jakob disease and related disorders in Europe, Australia, and Canada. Neurology. 2005;64(9):1586–1591.

7. Litzroth A, Cras P, De Vil B, Quoilin S. Overview and evaluation of 15 years of Creutzfeldt-Jakob disease surveillance in Belgium, 1998-2012. BMC Neurol. 2015;15(1):250. doi:10.1186/s12883-015-0507-x

8. Doi Y, Yokoyama T, Sakai M, Nakamura Y. Creutzfeldt-Jakob Disease Mortality in Japan, 1979-2004: Analysis of National Death Certificate Data. Journal of Epidemiology. 2007;17(4):133–139. doi:10.2188/jea.17.133

9. Stehmann C, Senesi M, Lewis V, et al. Creutzfeldt-Jakob disease surveillance in Australia: update to 31 December 2018. Communicable diseases intelligence (2018). 2019;43. http://152.91.69.116/internet/main/publishing.nsf/Content/75F30C0D2C126CAECA2583940015EDE3/$File/creutzfeldt_jakob_disease_surveillance_in_australia_update_to_31_december_2018.pdf

10. Stratton E, Ricketts MN, Gully PR. The epidemiology of Creutzfeldt-Jakob disease in Canada: a review of mortality data. Emerg Infect Dis. 1997;3(1):63–64.

11. Maddox RA, Person MK, Blevins JE, et al. Prion disease incidence in the United States: 2003–2015. Neurology. 2020;94(2):e153–e157. doi:10.1212/WNL.0000000000008680

12. HCUP NIS Description of Data Elements: NIS_STRATUM - Stratum Used to Post-Stratify Hospital. Agency for Healthcare Research and Quality; 2008. Accessed November 11, 2021. https://www.hcup-us.ahrq.gov/db/vars/nis_stratum/nisnote.jsp

13. Morris NA, Chatterjee A, Adejumo OL, et al. The Risk of Takotsubo Cardiomyopathy in Acute Neurological Disease. Neurocrit Care. 2019;30(1):171–176. doi:10.1007/s12028-018-0591-z

14. Holman RC, Belay ED, Christensen KY, et al. Human Prion Diseases in the United States. PLOS ONE. 2010;5(1). doi:10.1371/journal.pone.0008521

15. Holman RC, Khan AS, Kent J, Strine TW, Schonberger LB. Epidemiology of Creutzfeldt-Jakob Disease in the United States, 1979–1990: Analysis of National Mortality Data. Neuroepidemiology. 1995;14(4):174–181. doi:10.1159/000109793

16. Pape WJ, Forster JE, Anderson CA, Bosque P, Miller MW. Human Prion Disease and Relative Risk Associated with Chronic Wasting Disease. Emerg Infect Dis. 2006;12(10):1527–1535. doi:10.3201/eid1210.060019

17. Nakamura Y, Watanabe M, Nagoshi K, Yamada M, Mizusawa H. Geographic Difference of Mortality of Creutzfeldt-Jakob Disease in Japan. Journal of Epidemiology. 2007;17(1):19-doi:10.2188/jea.17.19

18. StataCorp. Stata Statistical Software: Release 15. Published online 2017.

19. U.S. Census Bureau QuickFacts: United States. Accessed January 1, 2022. https://www.census.gov/quickfacts/fact/table/US/RHI725221#RHI725219

20. Joinpoint Regression Program. Published online April 2022.

21. Occurrence and Transmission | Creutzfeldt-Jakob Disease, Classic (CJD) | Prion Disease | CDC. Published May 8, 2019. Accessed September 6, 2020. https://www.cdc.gov/prions/cjd/occurrence-transmission.html

22. ElSaadany S, Semenciw R, Ricketts M, Mao Y, Giulivi A. Epidemiological study of Creutzfeldt-Jakob disease death certificates in Canada, 1979–2001. Neuroepidemiology. 2005;24(1-2):15–21.

23. Will R, Alperovitch A, Poser S, et al. Descriptive epidemiology of Creutzfeldt-Jakob disease in six european countries, 1993–1995. Annals of neurology. 1998;43(6):763–767.

24. Barash JA, Desai RA, Patwa HS. Veterans Health Administration Information Systems as a Resource for Rare Disorders Research: Creutzfeldt–Jakob Disease as a Paradigm. Mil Med. 2012;177(11):1343–1347. doi:10.7205/MILMED-D-12-00198

25. Barash JA, West JK, DeMaria A. Accuracy of administrative diagnostic data for pathologically confirmed cases of Creutzfeldt-Jakob disease in Massachusetts, 2000-2008. American Journal of Infection Control. 2014;42(6):659–664. doi:10.1016/j.ajic.2014.02.002

26. Collins S, Boyd A, Lee JS, et al. Creutzfeldt–Jakob disease in Australia 1970–1999. Neurology. 2002;59(9):1365–1371. doi:10.1212/01.WNL.0000031793.11602.8C

27. Sikorska B, Liberski PP. Human Prion Diseases: From Kuru to Variant Creutzfeldt-Jakob Disease. In: Harris JR, ed. Protein Aggregation and Fibrillogenesis in Cerebral and Systemic Amyloid Disease. Subcellular Biochemistry. Springer Netherlands; 2012:457–496. doi:10.1007/978-94-007-5416-4_17

28. Young GS, Geschwind MD, Fischbein NJ, et al. Diffusion-Weighted and Fluid-Attenuated Inversion Recovery Imaging in Creutzfeldt-Jakob Disease: High Sensitivity and Specificity for Diagnosis. American Journal of Neuroradiology. 2005;26(6):1551–1562.

29. Iwasaki Y. Creutzfeldt-Jakob disease. Neuropathology. 2017;37(2):174–188. doi:10.1111/neup.12355

